# Optimizing clinical dosing of combination broadly neutralizing antibodies for HIV prevention

**DOI:** 10.1101/2021.07.26.21261134

**Authors:** Bryan T. Mayer, Allan C. deCamp, Yunda Huang, Joshua T. Schiffer, Raphael Gottardo, Peter B. Gilbert, Daniel B. Reeves

## Abstract

Broadly neutralizing antibodies are promising agents to prevent HIV infection and achieve HIV remission without antiretroviral therapy (ART). As learned from effective ART, HIV viral diversity necessitates combination antibody cocktails. Here, we demonstrate how to optimally choose the ratio within combinations based on the constraint of a total dose size. Optimization in terms of prevention efficacy outcome requires a model that synthesizes 1) antibody pharmacokinetics (PK), 2) a mapping between concentration and neutralization against a genetically diverse pathogen (e.g., distributions of viral IC50 or IC80), 3) a protection correlate to translate *in vitro* potency to clinical protection, and 4) an *in vivo* interaction model for how drugs work together. We find that there is not a general solution, and the optimal dose ratio likely will be different if antibodies cooperate versus if both products must be simultaneously present. Optimization requires trade-offs between potency and longevity; using an *in silico* case-study, we show a cocktail can outperform a bi-specific antibody (a single drug with 2 merged antibodies) with superior potency but worse longevity. In another practical case study, we perform a triple antibody optimization of VRC07, 3BNC117, and 10-1074 bNAb variants using empirical PK and a pre-clinical correlate of protection derived from animal challenge studies. Here, a 2:1:1 dose emphasizing VRC07 would optimally balance protection while achieving practical dosing and given conservative judgements about prior data. Our approach can be immediately applied to optimize the next generation of combination antibody prevention and cure studies.

## Introduction

Broadly neutralizing antibodies (bNAbs) are powerful agents that may become crucial for next generation HIV prevention^1^. Their utility is strengthened by their generally long half-lives compared to small molecule drugs, as well as the eventual promise of inducing bNAb production by vaccination^2,3^.

The recent antibody mediated prevention (AMP) studies directly tested the hypothesis that the VRC01 bNAb could prevent HIV acquisition^4,5^. While the study found no significant overall prevention efficacy, once HIV-1 Envelope pseudoviruses were made based on viral sequences from trial participants who acquired HIV-1 infection, it emerged that viruses acquired by placebo recipients were more sensitive to neutralization by VRC01 than viruses acquired by VRC01 recipients. The prevention efficacy against sensitive viruses (sensitive was defined as IC80 < 1 µg/ml) was estimated at 75.4% (95% confidence interval 45.5 to 88.9%). Less sensitive variants comparatively infected placebo and control recipients^5^. Indeed, the diversity of globally circulating strains^6^ remains beyond the breadth of any current bNAb. As with antiretroviral treatment (ART) and pre-exposure prophylaxis (PreP), combinations of products are likely needed^7–9^.

*In vitro* combination bNAb potency has been studied and modeled previously^10,11^. Here, we extend these pharmacodynamic (PD) models to incorporate *in vivo* concentrations over time (pharmacokinetics, PK) with multiple bNAb administrations to establish a PKPD framework for clinical design focusing on how to ration the dose of each bNAb. A particularly important component of our modeling is that we allow bNAb concentrations to vary and distinguish *in vitro* and *in vivo* potency^12^. As neutralization markers that best predict prevention efficacy (PE) are still under investigation, we consider flexible choices of the PKPD outcomes to be optimized, and also show that many outcomes are co-optimized. We apply our framework to 2 realistic *in silico* case studies. The first is a comparison of two antibodies against a bi-specific antibody^13,14^, a synthesized combination of the two “parental” antibodies. We assume the bi-specific gains potency through the combination, but loses longevity, clearing with the faster of the two parental antibody half-lives. The second study models a three-drug combination of clinical candidate bNAbs (VRC07-523-LS^15^, 10-1074 & 3BNC117^16^) and applies a protection correlate—protection predicted by neutralization titer—derived from non-human primate challenge studies^17^.

Our framework is designed to answer a crucial design consideration for these future studies: what is the optimal ratio of multiple antibodies to deliver in a single dose of a fixed size? We show the optimal ratio can depend on any and all inputs and assumptions -- precluding a one-size-fits-all solution. Instead, we provide a framework and a publicly available tool to determine the best dose plan given the specific antibodies, existing information about their interaction *in vivo*, and the PKPD outcome marker of interest for a proposed study. As more is known about each of these components, the model framework will rely on less uncertainty and become more predictive.

## Results

We previously integrated pharmacokinetic (PK) and multi-strain pharmacodynamic (PD) models to determine longitudinally varying potency of VRC01, a broadly neutralizing antibody (bNAb), to simulate prevention trials and predict strain coverage^18,19^. Because of HIV genetic diversity, it is essential to consider the distribution of potencies against the diverse population of viral strains. We now extend this framework to model multiple bNAbs, where integrating the combination PD models with PK adds several layers of complexity (**Fig 1**).

**Fig 1.**
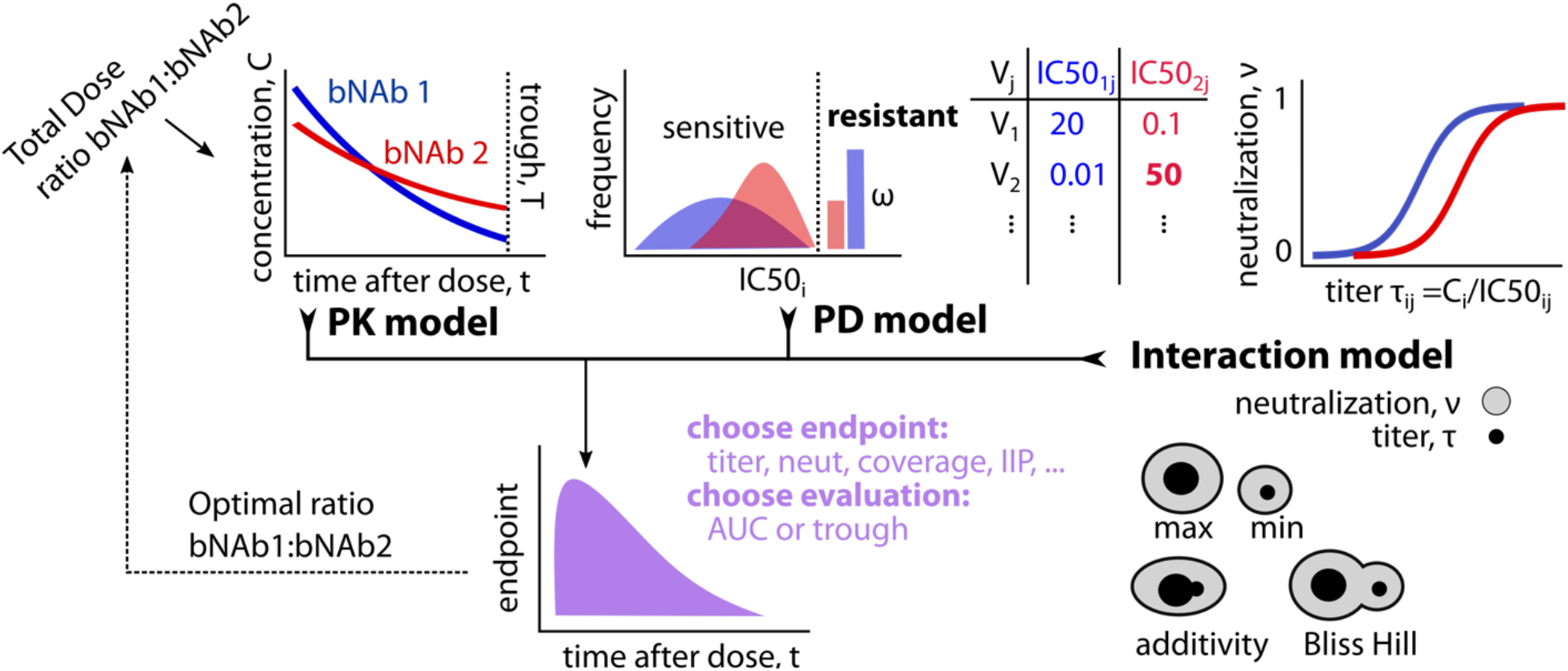
PKPD model schematic for optimizing combination treatment against a genetically diverse pathogen. The model incorporates: pharmacokinetics (PK), pharmacodynamics (PD), and interactions between antibodies. The PK describes antibody concentrations over time after administration. The PD model describes the distribution of neutralization potencies for each antibody against a variety of viral strains (quantified here by IC50ij, the level of the i-th drug needed to achieve 50% neutralization of the j-th viral strain). We also allow some fraction ω of strains to be completely resistant. Then, from titer, or the ratio of concentration to IC50 of each antibody against a certain strain, we define neutralization using a logistic function, which defines the proportion (0-1 scale) of viruses that are neutralized. There are four functional PD interaction scenarios. The first two are heuristic taking either the worst (minimum) the best (maximum) titer or neutralization between two products. The other two are mechanistic combinations described by independently combining titers (additivity) or neutralization (Bliss Hill). Then, depending on the PKPD outcome measure of interest (for example titer, neutralization, coverage defined below) and when that measure should be evaluated (throughout the study = AUC, at the low point = trough), we identify the optimal ratio of the antibodies to be included in the initial dose.

### Pharmacokinetics (PK) for bNAb levels

The first component of the PKPD framework is the PK, describing concentrations of each antibody *i* over time, *t*: *C*_*i*_(**θ**_*i*_, *t, d*_*i*_) where **θ**_*i*_ are the bNAb specific PK parameters and *d*_*i*_ is the initial dose (PK model in **Fig 1**). Individual initial dosing for each bNAb is then constrained by a total dose (*D* = Σ_*i*_ *d*_*i*_). For simplicity, we assume a population-level fixed total dose and independent models of PK for multiple bNAbs (denoted *C*_*i*_(*t*) from here on). The model could be extended to implement individual-specific total dosing (e.g., bodyweight-adjusted) and joint, dependent models.

### Pharmacodynamics (PD) for bNAb potency

Two pharmacodynamic (PD) quantities are often used to discuss neutralization given concentration: 50% inhibitory dose or dilution neutralization titer (ID50 Titer) and percent neutralization. Both quantities incorporate concentration and 50% inhibitory concentration (IC50) measurements across a panel of viruses (PD model in **Fig 1**).

Experimental neutralization titer (ID50), *τ*_*ij*_(*t*), is a common measurement arising from titrated neutralization experiments. In practice, experimental ID50 represents a dilution factor applied to sera containing antibodies that reduces *in vitro* neutralization to 50%. Titer, and the similarly derived ID80, are important immunological endpoints that are proven correlates of protection^4,17^. Experimental titer can be theoretically predicted from the ratio of *i*-th drug concentration to *j*-th virus IC50 as

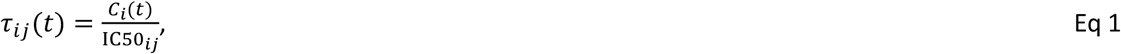

a relationship that has been empirically confirmed for single bNAbs^19,20^. As a potency measure, titer expresses the fold-relationship between concentration and viral IC50 as a measure of ‘protection’ against that virus.

Experimental *in vitro* neutralization for a single bNAb against a virus is also theoretically related to the titer (**Fig 1**). Neutralization, on a 0-100% scale, has the mechanistic interpretation of the fraction of blocked cellular infection events by the *j*-th virus, or “% neutralization”. Titer and neutralization, *ν*, can be related through the logistic Hill function (or median-effect equation) as follows

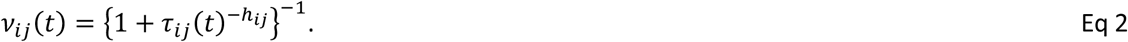

Neutralization requires an additional parameter, the ‘Hill coefficient’ *h*_*ij*_, that describes the steepness of the neutralization curve. Through Eq 2, any generalized titer (e.g., ID80, ID99) can be predicted from the ID50 titer and a given Hill slope, where the Hill slope can be estimated from IC50 and IC80 measurements (see **Supplementary Information**). Using the CATNAP database^21^ of IC50 and IC80 neutralization estimates for HIV virus/antibody combinations, we estimated the distribution of Hill slopes and generally found values near 1 (See Methods and **Supplementary Figure 1**). Henceforth in our analysis, and consistent with previous measurements^18^, we set *h*_*ij*_ = 1 and it is dropped from equations. Under this assumption, the IC80 is theoretically predicted to be 4-fold higher than the IC50, and, subsequently, the ID80 is predicted to be 4-fold lower than the ID50 for single bNAb and virus combinations (see **Supplementary Information** for more details).

### bNAb interaction models

For bNAb combinations, we considered 4 PD interaction models. The first, Bliss-Hill independence (BH), is the best-case multiplicative interaction where bNAbs cover missing breadth of one another and co-neutralize strains, i.e., virions must escape independent binding events from each antibody. BH is encouragingly observed from *in vitro* studies^10,22^. We also consider weaker cooperation with the additivity interaction model, where antibody effects are combined via mass action^10^; i.e., the total titer is sum of individual titers. Finally, maximum and minimum models assume that the more or less potent antibody for each strain operates as a single product. The maximum interaction potentially represents a scenario where only the most potent bNAb neutralizes a given virus; however, outcome deviations between the maximum and the BH or additivity model also highlight where interactions improve neutralization due to combined coverage. On the other hand, the minimum model is mechanistically unrealistic but provides a boundary for the worst-case scenario where the combination regimen is only as strong as its weakest link, specifically penalizing poor combined coverage of viruses.

The interaction models are mathematically summarized in **Table 1** and all derivations of combinations titers are included in the **Supplementary Information**. We extend interactions to include synergy in the bi-specific antibody case study, but do not consider antagonism among clinically viable bNAb combinations here.

**Table 1.**
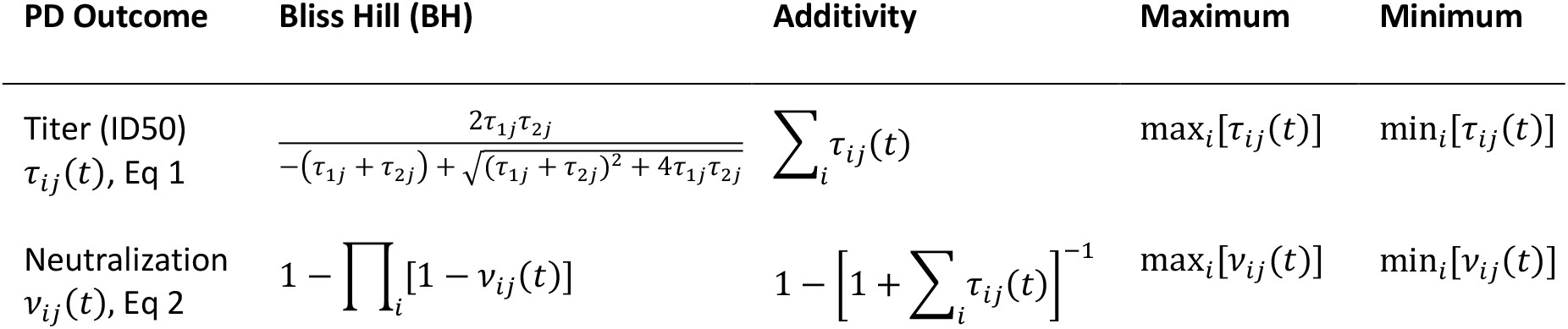
Summary of equations for PD interaction models relating bNAb (*i*) to virus (*j*). Formula for Bliss-Hill ID50 illustrated for 2-bNAb combinations only.

Other options exist to quantify antibody potency, including instantaneous inhibitory potential (IIP^23^), the log-fold reduction in virus infectivity at a given concentration, which linearizes high neutralization on the log-scale (e.g., 99% neutralization -> IIP of 2, 99.9% -> 3) in the important range for ART efficacy^23^.

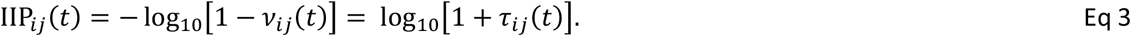

A generalized version of IIP when *h*_*ij*_ ≠ 1 is described in the **Supplementary Information**.

Alternatively, the potency of an antibody combination can be quantified by its “viral coverage”: what fraction of viral strains are above a specified threshold value. Fundamentally, the neutralization measurement is dichotomized for a given bNAb/virus combination, i.e., the virus is neutralized or not based on some measurement threshold. For example, for *n* strains and a neutralization threshold *ν*^*^, we define the neutralization coverage fraction 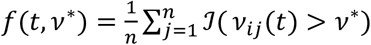 where 𝒥 is the indicator function equal to 1 if the inequality holds and 0 otherwise.

### Mathematical model for optimizing antibody combination doses

Finally, we summarize these measurements of potency over time, which we collectively term PKPD outcomes. We consider PKPD outcomes at trough (pre-specified final time) or throughout time (area under the curve, AUC) (**Fig 1**).

In practice, for a specified antibody combination, we obtain their PK parameters and the best estimate of their distribution of IC50s to a relevant panel of circulating viruses. We can then choose an interaction model and specify an outcome that we want to optimize. From this we uniquely determine the optimal ratio of the antibodies. Potential combinations of bNAbs—varying by their input PK and PD profiles—can then also be evaluated and compared via mathematical PKPD simulations at the optimal dosing ratios, which may be combination-dependent, as illustrated in the *in silico* studies below.

### Global sensitivity analysis

Across a range of theoretical 2-bNAb combination studies, we performed a global sensitivity analysis varying all input PKPD model parameters (**Fig 1**) to assess correlation between all PKPD outcomes and optimized dosing ratios (see **Methods**). Briefly, we varied one-compartment exponential PK models for each antibody summarized by their half-life *hl*_*i*_. = ln 2/*k*_*i*_. The PK model used a one compartment model with fixed volume of distribution (3 L)^32,33^. One bNAb was simulated to always have equivalent or better half-life than the other to avoid redundancy. We chose a log-normal distribution for IC50s for each bNAb parameterized by its mean *μ*_*i*_ and standard deviation *σ*_*i*_ on the log10 scale, also allowing for a fraction *ω*_*i*_ that are completely resistant (infinite IC50). We also varied the ratio of doses *r* and the total dose *D*. The ranges explored for each sensitivity analysis parameter are collected in **Table 2**.

**Table 2.** Parameter settings for PKPD sensitivity analysis for combining 2 bNAbs.

| Parameter | Sensitivity analysis values |
| --- | --- |
| Initial dose (mg) | {150, 300, 600, 1200, 2400} |
| Half-life (days) | {7, 28, 42, 84} |
| Total simulated viruses | 500 |
| % viral resistance | {67, 33, 0} |
| Mean log10 IC50 ($\mu\text{g/mL}$) | {-3, -2, -1} |
| SD log10 IC50 ($\mu\text{g/mL}$) | {0.25, 0.5, 1} |

### Publicly available tool for ratio optimization

Any individual simulation from the results can be generated using the following R shiny app: https://bnabpkpd.fredhutch.org.

### PKPD outcomes cluster into categories

Using global sensitivity analysis output, we calculated Spearman correlations among all endpoints at trough (**Fig 2A**). By hierarchical clustering, we determined six main categories of outcomes (**Fig 2A**): All models with the minimum interaction (i.e., worst-case bNAb penalizing lack of combination viral coverage) and raw titer (ID50) endpoints for the non-minimum interaction were quite distinct. The remaining outcomes were correlated but further categorized as neutralization, log10-transformed titer, coverage metrics (% of viruses neutralized > 99%), and IIP. Results were similar for AUC and trough, see **Supplementary Fig 2**, which indicates for a simple monotonic PK curve, the final value is representative of the entire time-course.

**Fig 2.**
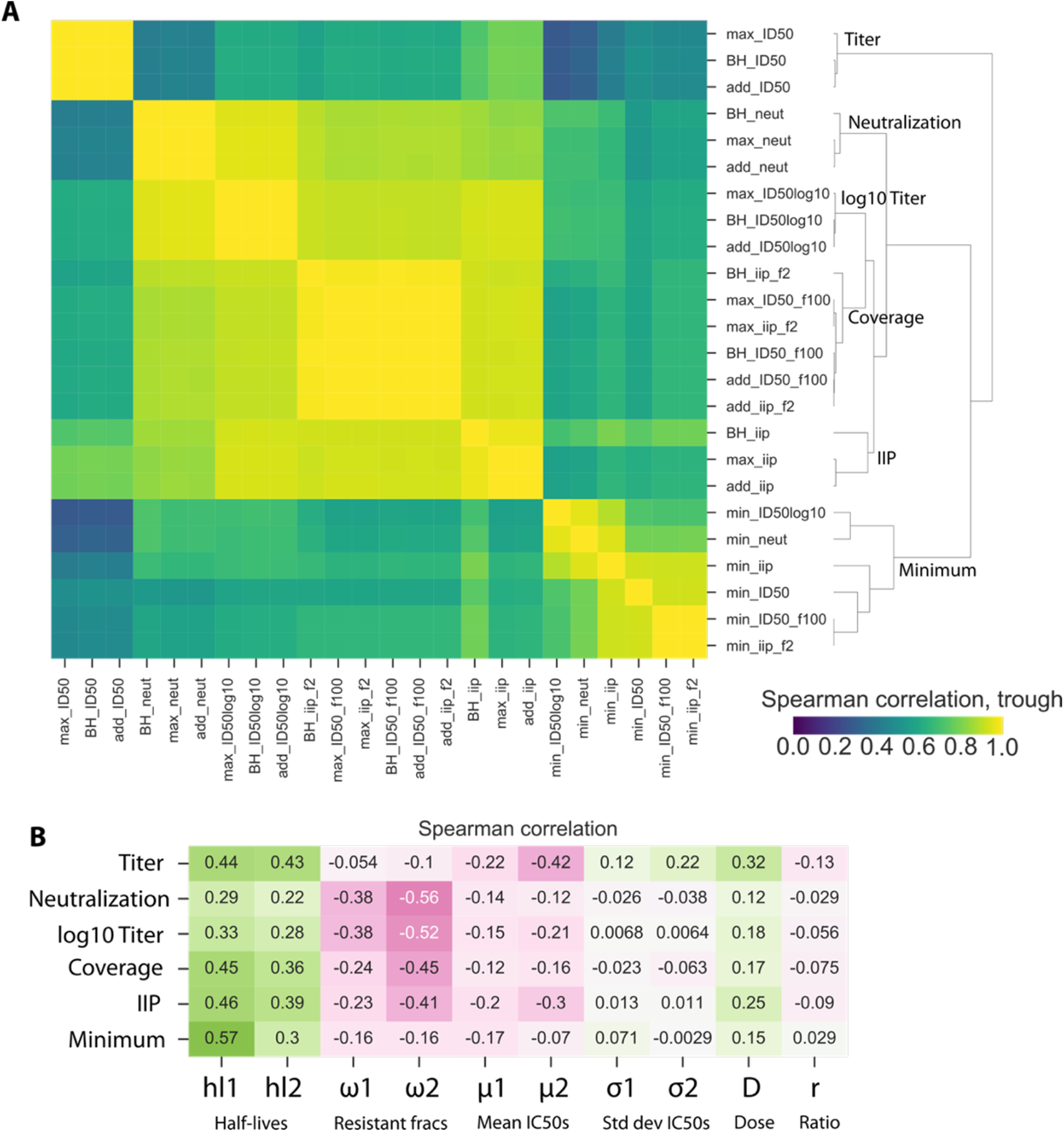
Correlations among PKPD outcomes and between model parameters and outcomes. We performed a global sensitivity analysis to simulate a two-drug combination therapy against a genetically diverse pathogen where the drug 1 had equivalent or worse half-life than drug 2. We simulated 24 outcomes at trough (results for AUC are similar), including ID50, log10 ID50, neutralization, instantaneous inhibitory potential (IIP), as well as the coverage fraction of pathogen strains having ID50>100 and IIP>2. A) Many of these outcomes are strongly correlated (yellow in heatmap). Moreover, these 24 outcomes cluster into approximately 6 distinct categories: see labels along dendrogram. Of the interaction models, only the minimum interaction separated into its own category while the others clustered together within a given outcome. B) Overall, we varied 10 model parameters (Table 2). By correlating (Spearman, green∼1, pink∼-1) to the 6 categories from panel A against all model parameters, we found that the categories were similarly sensitive to PK, while titer and minimum categories were less sensitive to resistance fractions. The ratio does not strongly predict any outcomes when all other parameters are varied, highlighting that there is no general solution to optimizing the ratio and it must be adjusted on a case-by-case basis.

### Correlations among PKPD outcomes and antibody features

We next explored the associations between a representative member of each outcome category and model parameters (**Fig 2B**). All categories were sensitive to PK (half-life), and generally more to the half-life of the shorter-lived bNAb (hl2). Increased resistance negatively correlated with the outcomes, particularly with neutralization, log10-transformed titer, coverage metrics, and IIP. Additionally, a stronger negative correlation was found with the resistant fraction for the bNAb with longer half-life – this pattern was weaker for mean IC50. Total dose correlated positively with all outcomes but was generally less influential than other model parameters. The ratio of antibodies did not strongly predict any outcome after accounting for variation in all other parameters, highlighting that there was no generally optimal ratio; optimization is determined on a case-by-case basis based on many antibody features.

### Sensitivity of the optimal ratio for each outcome

Next, for each parameter set, we determined the optimal ratio *r* for each outcome. **Fig 3A** shows an analogous clustering analysis to **Fig 2** but with correlations of the *optimal ratio* of each outcome across the inputs. Importantly, the same categories emerged such that correlations among all outcomes agreed generally with correlations among optimal ratios. In particular, the optimal ratio for minimum not only appears distinct from the other categories but is often negatively correlated to the others. This suggests that optimizing for minimum interaction (i.e., maintaining consistent combination coverage) may require a very different ratio. For the other interactions, once an outcome is selected, the optimal ratios generally agree among maximum, additive, and Bliss-Hill interaction models.

**Fig 3.**
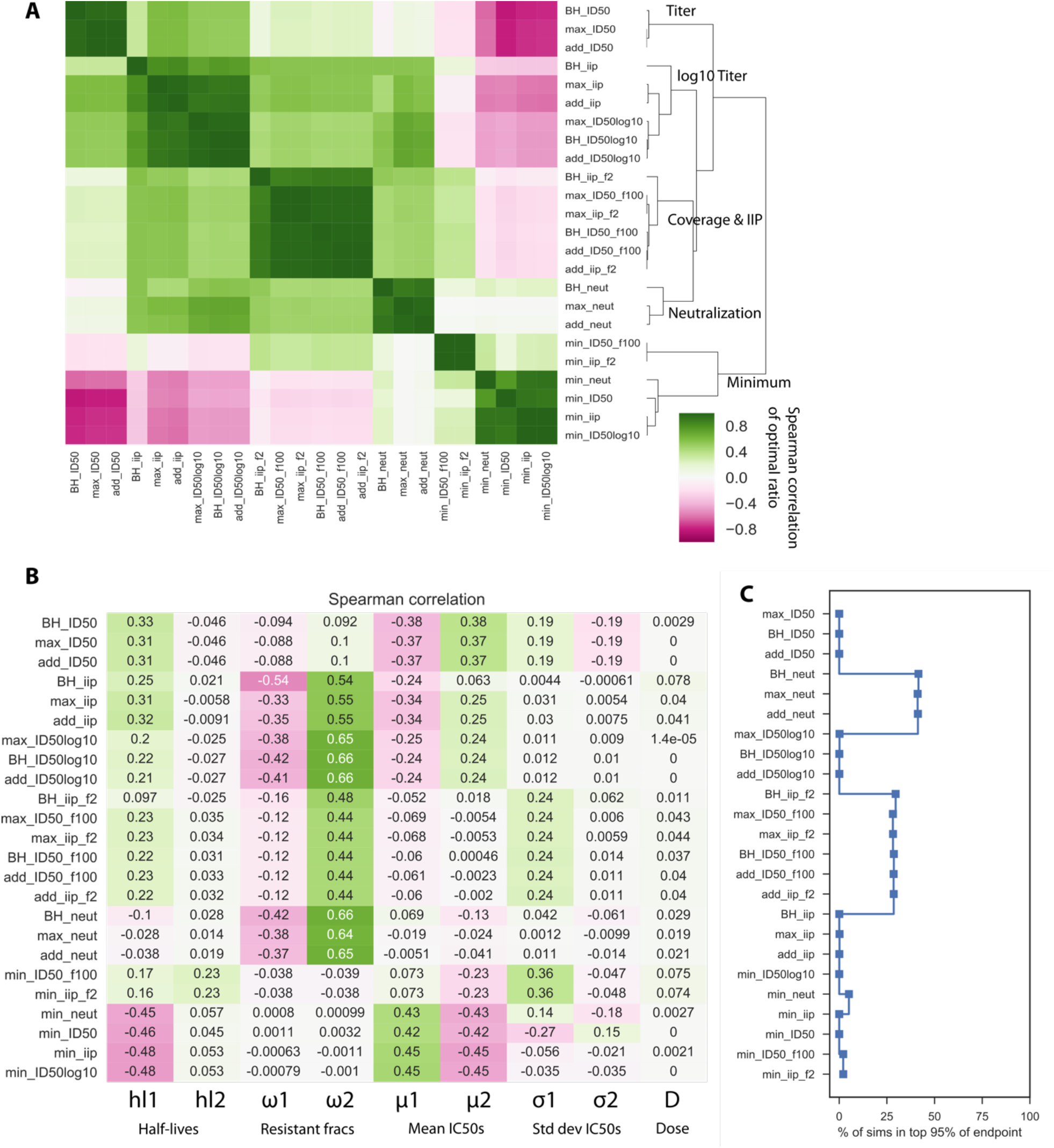
Sensitivity of the optimal ratio to PKPD outcome choices and antibody features. From the global sensitivity analysis, we calculated the optimal ratio for each parameter set and each outcome. (A) We repeated the clustering analysis to determine how the optimal ratio clusters by outcome. In this setting coverage and IIP cluster together leading to 5 (rather than 6) categories, but the others remain the same from the prior analysis in **Fig 2**. B) Certain variables drive optimization for different outcomes. For example, the mean IC50 are most influential on optimizing ratios using minimum neutralization and titer outcomes, while the fraction resistant is most influential for the remaining outcomes. C) Across all simulations, we quantified the sensitivity of the optimal ratio of each outcome by calculating the fraction of parameter sets in which the outcome was within 95% of its optimal value. For the parameter ranges considered, some of these outcomes were not enhanced greatly by perfect optimization, but outcomes clustered in the correlation showed similar sensitivity and some outcomes were particularly sensitive.

**Fig 3B** shows correlations among optimal ratios for each outcome and model parameters. Here directionality of correlation has additional meaning: positive and negative correlations imply less or more of the antibody with worse half-life, respectively. The sensitivity to PK and PD (resistance and mean IC50) followed the same pattern as in **Fig 2A**: all the outcomes showed some sensitivity to PK, titer and minimum interaction outcomes were sensitive to mean IC50, and the remaining were sensitive to resistance fractions. For ratio optimization, the PK sensitivity was specifically driven by the half-life of the shorter-lived bNAb.

We next sought to understand what is gained by using the optimal ratio as opposed to a more practical solution near the optimum. Therefore, we measured how many parameter combinations admitted an outcome within 95% of the outcome value achieved by the optimal ratio (**Fig 3C**). That is, if most simulations were within 95% of the optimum, it means the optimum is not substantially better. Indeed, for the parameter ranges we considered, some outcomes were not particularly sensitive to the choice of the optimal ratio such that other practical considerations could be promoted in a trial design. However, some outcomes were much more strongly affected by optimization (with fewer than 1/10^4^ runs being in the 95% optimal scenario) including IIP. So, although there are cases of insensitive systems (e.g., two poor products, two highly effective products), this reinforces that optimization should be case-specific.

### Dual parental antibodies outperformed bispecific product without synergy enhancement

Bi-specific antibodies, synthesized combinations of two antibodies into one product, appear *in vitro* to exhibit superior neutralization compared to their parental components^13,14^. However, these experiments are not inclusive of *in vivo* pharmacokinetics. Bi-specific antibody clearance may be determined based on the clearance kinetics of either parental component. If clearance characteristics are comparable to the slower of the two parental antibodies, then the bi-specific combination is clearly advantageous. Because bi-specific PK is not well studied, we tested the non-trivial scenario in which a bi-specific inherits the worse (faster) parental half-life. We investigated a realistic design administering 300 or 1200 mg of antibodies and 3-month administration window. We assumed one parental antibody had a 3-month half-life equivalent to the trough time but with worse PD than a superiorly potent bNAb. However, the more potent bNAb was given a poorer half-life of 1 week (i.e., equivalent to 1/12 of trough window). We evaluated the theoretical study efficacies using the following PKPD outcomes: a continuous outcome (mean IIP) and a coverage outcome (% viruses IIP>2).

Compared to the combination therapy, the superior potency of the bi-specific antibody is not necessarily sufficient to account for a poor PK profile. Across doses and interaction models, we consistently found that the optimal combination therapy was more efficacious than the bi-specific for both AUC and trough (**Fig 4**). At trough, where half-life had higher influence, the parental with better half-life but worse PD alone outperforms the bi-specific particularly at lower doses.

**Fig 4.**
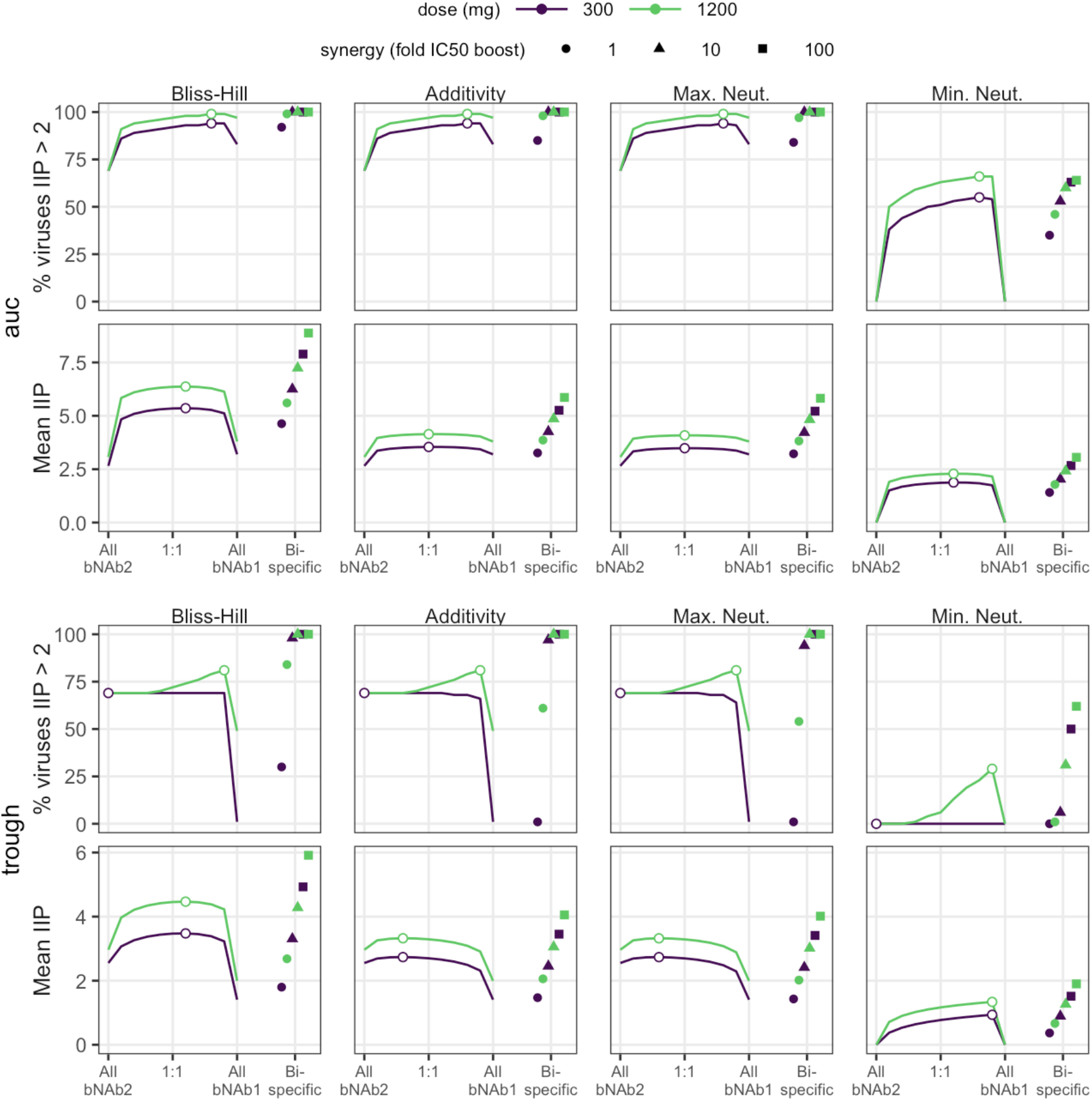
Optimizing 2 bNAb combination therapy in comparison to bi-specific therapy with the same bNAbs. *Combination antibody results for AUC (top) and trough (bottom) suggest that trough is slightly more sensitive to ratio (see curvature of outcome surface and change from optimal ratio denoted by open dot). In general, a single bi-specific bNAb will perform worse than combination therapy if it has the best neutralization potential of both parental lineages under a common interaction model but inherits the faster clearance kinetics. However, if synergetic binding occurs, enhancing the bi-specific potency by 10-fold (see* ***Methods****), it is similar or outperforms the optimal combination for all outcomes and doses*. “All bNAb1” and “All bNAb2” on the x-axis correspond to 100% dosing of the second bNAb product.

**Fig 5.**
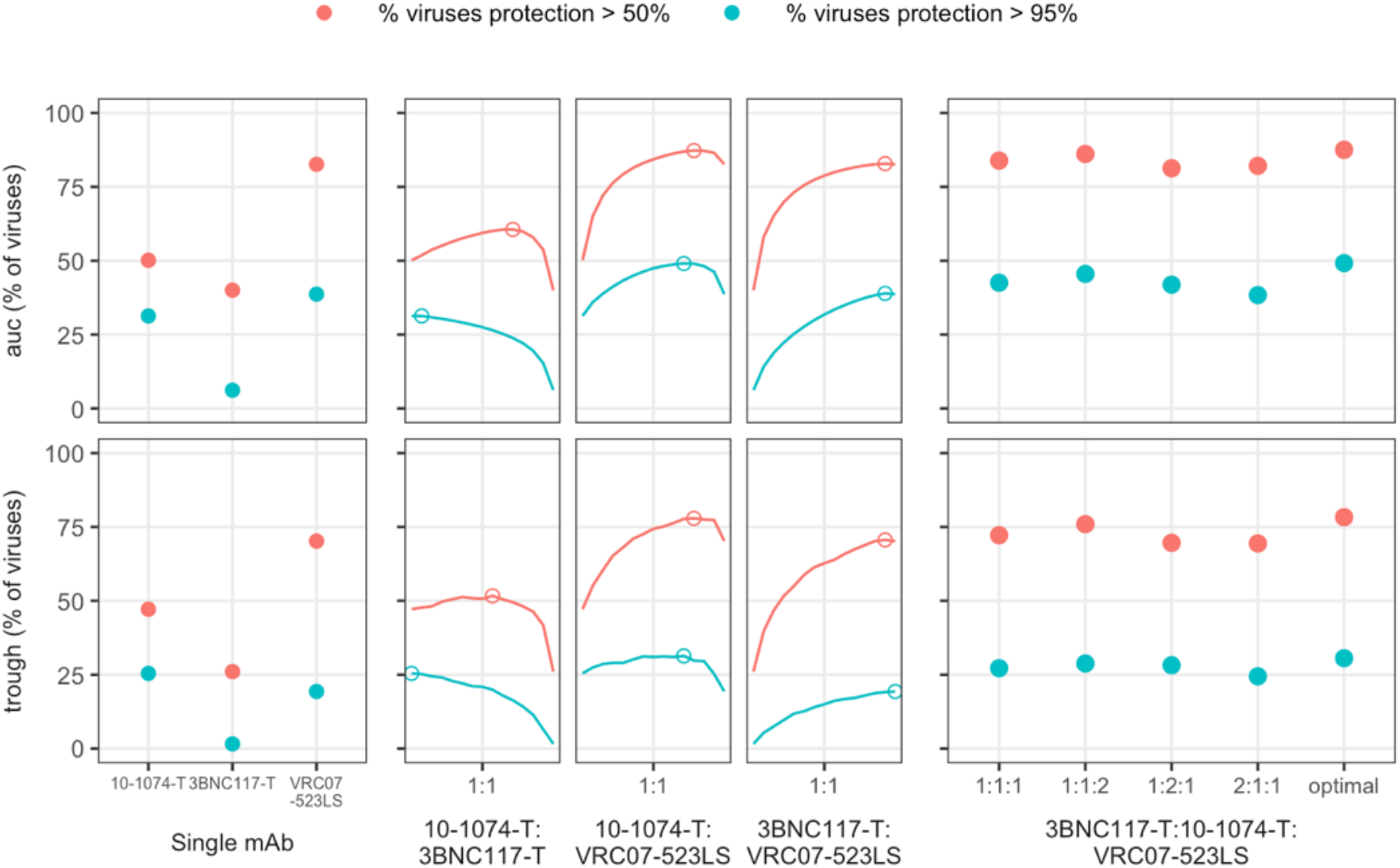
Additional enhancement after optimization of 3-drug therapy. Using 3 well known anti-HIV broadly neutralizing antibodies, we performed an analysis comparing the percent of viruses at more than 50% and 95% neutralization level for the bNAbs individually, in 1:1 combination, and in triplicate as 1:1:1, 1:1:2, 1:2:1, 2:1:1, and the optimal combination (see **Table S2**). Enhancement over the best single bNAb (VRC07-523-LS) is generated through combinations when evaluating the percent of the viruses neutralized at a 95% level. However, triple drug therapy does not meaningfully enhance over optimized 2-drug therapy levels, even when completely optimized. Indeed, a 1:1:1 3 drug therapy is outperformed by the optimized 2-drug therapy, highlighting the need to carefully perform case-studies for any optimization scenario.

Given this finding, we tested how much additional synergy (as a factor multiplying the bi-specific potency through reduced IC50, see **Methods**) could rescue the bi-specific performance and make it comparable to the parental combination. Synergy has been observed for bi-specifics because binding of one antibody arm can facilitate the second to bind^24^. Using synergy models, the bi-specific outperformed the optimized combined administration when the synergy factor exceeded 10-fold under common interaction models (**Fig 4**).

### Incorporating empirical protection correlates in clinical design

To perform a realistic optimization of a clinical trial, we consider deviations from *in vitro* potency that may be relevant for *in vivo* protection. For example, non-human primate HIV challenge studies suggest that a bNAb titer of approximately 100 achieved 50% protection: i.e., serum antibody concentrations need to be 100-fold higher than *in vitro* IC50 to elicit 50% protection *in vivo*^17^. We define the fold-increase as a “potency reduction factor”^18^, *ρ*, and henceforth translate *in vitro* potency to *in vivo* protection by scaling the titer input. We have *in vivo* neutralization and IIP then,

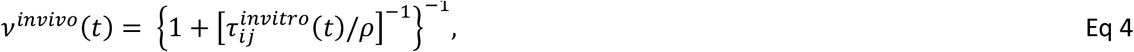

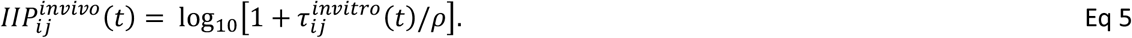

such that no change from *in vitro* measured titer occurs when *ρ* = 1 and a potency reduction of 100-fold means *ρ* = 100. Mechanistically, this formulation suggests that the overestimated protection *in vivo* is due to either (or both) underestimation of the potency due to some biological factors (e.g. coagulation or anti-antibody elements) or overestimation of bNAb concentration at the site of exposure.

The reduction factor can be derived from assessing actual protection at the given experimental titers, either through NHP challenge or using protection efficacy (PE) estimated from clinical study. Alternatively, if multiple protection estimates for varying titers, the titer vs. protection dose-response relationships can also be structurally varied; for example, we employed a 5-parameter logistic model on the NHP protection data for the following case study (see **Methods**).

For combination bNAbs, the experimental titer will represent neutralization in sera with a combined concentration of antibody. Whether a potency reduction factor is applied to the combination titer or to the individual titers prior to the interaction is specifically consequential for the Bliss-Hill interaction model, but not the other interaction models. Briefly, applying the factor to the Bliss-Hill combination titer model may be overly conservative, underestimating the protection because experimental titer does not uniquely predict Bliss-Hill neutralization (**Supplementary Figure 3**; see **Supplementary Information** for further discussion). We suggest applying the potency factor or protection model to each bNAb individually, calculating their individual protection estimate, then applying the Bliss-Hill interaction model (i.e., at the event-level) as described in the following case study.

### Using empirical protection correlates in a 3-bNAb optimization

We gathered several independent data sets to model a 12-week trial with a 600 mg subcutaneous dose of 3 state-of-the-art broadly neutralizing antibodies (3BNC117-T, 10-1074-T, VRC07-523-LS; -T denotes theoretical variant with extended half-life). For this example, we used an empirical protection estimates based on titer from the Pegu *et al*. NHP meta-analysis^17^ with the primary PKPD outcome of viral coverage at 50% and 95% protection thresholds. For more details on the input PK and PD for these analytes, see **Methods** and **Supplementary Figure 4**. In this illustrative example, we do not consider clade-specific profiles nor account for interference potentially due to 3BNC117 and VRC07-523-LS targeting the same epitope (CD4-bs). Next, we tested all double and triple combinations varying the dosing ratios. In a clinical setting, it is unlikely that complicated dosing ratios would be of practical consideration (e.g., 98:13:3). Thus, for the 3-bNAb combination, we considered simple ratio designs: an even dose split (denoted 1:1:1) or any 50%:25%:25% combination (denoted 2:1:1 or similar). We compared this to the theoretical optimum to ensure they were reasonably close to the optimal design.

Overall, all triple drug combinations predicted a protection level above 95% for roughly 25% of viruses at trough. Likewise, protection levels were above 50% for roughly 80% of viruses over the study (AUC). It was clear that VRC07-523LS was the best single antibody, and the optimal dosing ratio generally contained >60% of VRC07-523-LS (see **Table S2**). Subsequently, the triple combination with 1:1:2 level of VRC07-523LS was not much worse than optimal. Moreover, the optimal 2-drug combination without 3BNC117-T was nearly as effective as the optimal 3 drug therapy (which dosed at <10% of 3BNC117-T) potentially due to general lower potency of 3BNC117 or the overlap in epitope targeting with VRC07-523-LS resulting in redundant viral coverage in the database. Still, given our necessarily incomplete data on circulating strains, we would suggest using this 3-drug therapy at a 1:1:2 design to balance simplicity and protection for this example.

## Discussion

Combination administration of broadly neutralizing antibodies is likely to form a key component of future studies of HIV prevention^1,4,14,25^. While antibody neutralization is essential, accurate balancing of antibody dosing requires modeling both neutralization and concentration levels over time. Our approach here addresses this critical unmet need.

Our analysis highlights that several types of data for each antibody in a combination modality must all be considered to optimize dosing rationally. These include 1) the input potency data relating each bNAb to existing *in vitro* assays that test drug potency (i.e., IC50); 2) a translation to an *in vivo* protection metric = using a correlate derived from NHP meta-analysis; and 3) an understanding of drug interactions. The second step is crucial because in vitro IC50 measurements could underestimate *in vivo* efficacy^18,26^. While we illustrate use of a NHP correlate, human correlates may soon be derived from the AMP trials. The best correlate will likely need to be translational using readily available *in vitro* neutralization data for predictions^14,22^, which may be calculated using pseudoviruses (e.g., CATNAP database^21^) or breakthrough viruses in human infections^27^.

Here, we depict how to implement a titer protection correlate into clinical design for combinations. As illustrated in our practical case study design using the NHP challenge correlate, we derive a dose-response relationship for titer and protection from the NHP challenge studies, and then assess combinations using this empirical protection as the PKPD outcome via viral coverage. We also highlight that the protection estimates derived from single bNAb studies need to be carefully translated into combination bNAb target outcomes. Specifically for the Bliss-Hill interaction, a combination sera titer may correspond to different protection estimates depending on the underlying individual concentrations of each bNAb. We suggest determining antibody-level protection first then applying the BH interaction. This approach is also amenable if different potency reduction factors or dose-response protection relationships are bNAb-specific depending on target site.

Because there remains uncertainty regarding the optimal PKPD protection endpoints for bNAb combinations, our sensitivity analysis illustrates several main categories with similar properties. Moreover, we show certain features of antibodies (long half-lives, broad coverage, etc.) are particularly predictive of success, raising the possibility of using these results to inform endpoint selection based on coarse knowledge of circulating strains. While PK heterogeneity will affect endpoints and should be considered for optimizing trial design, the endpoints we tested exhibited much more sensitivity to the PD profile of the product. The optimal ratio of a two-drug therapy was shown to be strongly sensitivity to specifics of the combined antibody features. Thus, in combination with additional demographic considerations and population risk, we advocate for specific optimization for any trial rather than relying on general rules. Additionally, certain endpoints are more sensitive to optimal dosing than others, which can be considered in endpoint selection, or alternatively, if an endpoint is preferred which is not particularly sensitive, practical considerations about dosing could be prioritized over precise dose optimization as illustrated by our 3-bNAb combination example.

In our sensitivity analysis, we also find that PKPD outcome levels and optimal ratio are well correlated between the maximum, additivity, and BH interaction. This suggests that the design of the trial is not particularly sensitive to selecting the correct interaction model among these choices; however, the correct choice may still improve the accuracy of the predicted PKPD outcome. On the other hand, the minimum interaction formed a unique cluster of simulated endpoints and optimized compared to others. While the minimum interaction may be unrealistic, it explicitly penalizes designs that lack effective combination coverage. In practice, a trial optimization may evaluate both a minimum and Bliss-Hill interaction endpoints, allowing the minimum interaction to represent a worst-case scenario where there is no protection against viruses without sensitivity to at least two bNAbs.

Although most of our analysis concerns prevention studies, this framework is applicable to curative studies attempting to use bnAbs to prevent viral rebound after stopping ART^28,29^. The challenge in this setting is within-host diversity in the reservoir. Blocking a single founder during a transmission event appears easier than blocking repeated reactivations of diverse viral populations. Several studies have illustrated bnAbs can delay viral rebound^28–30^. However, levels required to prevent rebound remain hard to predict. In a cohort of 18 individuals receiving VRC01 infusion and ART cessation rebound occurred when plasma VRC01 levels were well above *in vitro* IC50s^29^.

Our analysis shows potential limitations around bi-specifics once PK is considered. Specifically, if the synthetized product clears faster, performance can be worse than an optimized combination therapy of the two parent products. The prevention benefits from bi-specific products thus rely on beneficial co-binding represented through some synergy, but these benefits may trade off with poorer half-life. Without synergy, bi-specifics are effectively a 2-fold concentration bonus, but neutralization is relatively insensitive on this scale, requiring input changes on the log-scale to either IC50s or concentrations. Of note, a powerful synergistic effect may allow these products to be potent at unmeasurable concentrations (i.e., below a typical limit of detection), which may not be tenable for study and practical use in a clinical setting. On the other hand, bi-specifics may be clinically preferable as they are a single product, and if the PK is at least half the dosing interval (or trough time), then our analysis suggests they theoretically perform comparably or better than combinations without consideration of synergy.

The three-drug optimization exercise illustrates that one potent antibody can determine the ability of combinations. Indeed, in this specific example, a two-drug therapy would have been nearly as good. However, in considering that viral panels are necessarily incomplete, we would err on the side of inclusivity to both widen breadth and account for uncertainty about escape mechanisms. In this example, adding the third and optimizing the triple-drug ratio is always beneficial to the two-drug combination, albeit minorly.

Going forward, our recommendation for designing therapeutic combinations for prevention or treatment of diverse pathogens is several fold: 1) choose outcomes based on expert opinions and given disagreements, assess whether these qualitative decisions are actually quantitatively in agreement; 2) consider multiple, distinct outcomes to evaluate a range of potential results; 3) optimize drug ratios for the specifics of component features; and 4) include subdominant levels of weaker antibodies to potentially cover holes in coverage not observable from incomplete preliminary data.

## Methods

### Code and data

All analysis were performed in R and Python. Simulations, data processing, and visualizations performed using R used the *tidyverse* package suite^31^. Sensitivity and cluster analysis of simulation results with subsequent visualizations were performed using the seaborn library in Python. All code will be available on GitHub.

### Estimation of Hill slope using CATNAP data

The Hill slope in the 2-parameter logistic Hill function (Eq 2) can be estimated from the IC50 and IC80 measurements (formula derived the **Supplementary Information**). We estimated the distribution of the neutralization Hill slope by performing this calculation across virus/antibody combinations available in the LANL CATNAP database^21^. To accommodate assay quantification limits that potentially vary across experimental study, we limited the analysis datasets to IC50 and IC80 values between 0.01 and 20 ug/mL, comprising 20,236 total combinations. Additionally, we grouped calculations within quartiles of input IC50 to assess whether Hill slopes vary by underlying viral sensitivity or measurement error that varies with the scale of IC50.

### Global sensitivity analysis

We performed ∼10,000 simulations over all combinations of parameters in Table 2 and calculated all PKPD outcomes. We chose a one-compartment exponential PK model with trough time 84 days for each bNAb: *C*_*i*_(*t*) = *C*_*i*_(0) exp(−*k*_*i*_*t*), and summarized the PK model with its half-life *hl*_*i*_ = In 2/*k*_*i*_. The PK model used a one compartment model with fixed volume of distribution (3 L)^32,33^. One bNAb was simulated to always have equivalent or better half-life than the other to avoid redundancy. We chose a log-normal distribution for IC50s for each bNAb parameterized by its mean *μ*_*i*_ and standard deviation *σ*_i_ on the log10 scale, also allowing for a fraction *ω*_i_ that are completely resistant (infinite IC50). We sampled 500 viruses per simulation. We then varied these parameters, along with the ratio of doses *r* and the total dose *D*. Then, we determined the optimal ratio as the ratio that maximized each PKPD outcome for all other parameter values across interaction models. To calculation IIP under Bliss-Hill, neutralization calculated for each bNAb and used as input, not titer (see **Supplementary Information**). Using the seaborn package in Python, we performed hierarchical clustering of Spearman correlations among outcomes and between parameters and outcomes.

### Comparison of bi-specific to parental antibodies

For the first parental bNAb, we chose a potent neutralizer (mean IC50 of 10^−3^ with 0% viral resistance) but with poor PK: elimination half-life equivalent to 1/12 of the administration period (i.e., 7-day half-life for an 84-day trough). For the second bNAb, we chose a more modest neutralizing profile (mean IC50 of 10^−2^ with higher variance and 33% viral resistance) but with excellent PK: elimination half-life equivalent to one administration period.

To model the bi-specific, we assumed the single molecule formulation means two parental products are given at the identical dose. We also assumed the clearance PK was determined by the faster of the two parental products. We additionally allowed for synergy, such that each antibody’s potency is improved by a factor *α*. This factor was assumed to be the same for all viral strains. Thus, following **Eq 3** and **Table 2**, the bi-specific IIP against a single virus *V*_*j*_ can be calculated for max, min, and additivity models, respectively

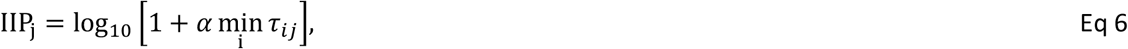

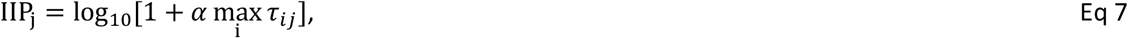

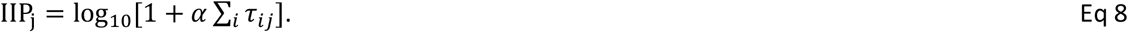

For Bliss-Hill interaction, the derivation from individual titers to a combination IIP is shown in the **Supplementary Information** (**Eqs S27** & **S29**) and then the bi-specific synergy was implemented as follows:

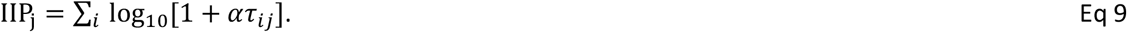

For comparing the combination and bi-specific therapies, we examined IIP and % viruses having IIP>2 (a surrogate of protection in nonhuman primate studies^17^) for AUC and trough. Calculations were based on 500 simulations as implemented for the global sensitivity analysis.

### Realistic clinical trial simulation

The full trial design contained a 12-week observation window and 600 mg total subcutaneous (SC) dosing with PK parameters established from clinical study (Table S1 and **Supplementary Fig 4A**). To boost performance of 3BNC117 and 10-1074, we artificially enhanced their half-lives by 3-fold to mimic an -LS variant (3BNC117-T and 10-1074-T). The distribution of *in vitro* neutralization against circulating strains was modeled using *in vitro* derived IC50s from 507 available common strains in the LANL CATNAP database^21^ (**Supplementary Fig 4B**).

We tested several models to map *in vivo* protection from *in vitro* neuralization. Pegu et al. developed a logistic regression model to predict protection probability from *in vitro* neutralization titer^17^. We use the output of their model at 50%, 75%, and 95% protection to test our model (**Supplementary Fig 4C**). Specifically, we employed the following approach: for a given bNAb (*i*) at a given concentration, we estimated *in vivo* protection (*p*) using neutralization titer (*τ*_*ij*_) against a virus (*j*). Using **Eq 3** and estimating a single parameter, the potency reduction, found *ρ*=1/91 and led to reasonable fits. However, a better fit was achieved using a 5-parameter logistic (5PL) model, a generalized dose-response type function with 5 parameters {*A, B, C, D, E*} and the form

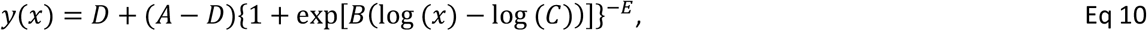

here mapping *in vitro* titer *x* = *τ*_*ij*_ and to *in vivo* protection *y* = *p*_*ij*_. We fixed *D* = 0 and *A* = 1 so that protection ranges from 0-1. The remaining 3 parameters were estimated as *B* = −1.84, *C* = 257, and *E* = 0.338. The best fit of the potency reduction model and the 5PL model are compared in **Supplementary Fig 4C**.

We then illustrate predictions of the 5PL model for each bNAb via % viral coverage at *in vitro* neutralization >50% compared to *in vivo* protection >50 and >95% in **Supplementary Fig 4D**. Using this model of protection, we then calculated combined protection across the administered bNAbs (*b* is the number of antibodies considered) assuming independence similar to Bliss-Hill:

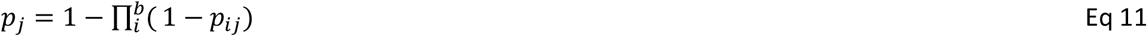

We then defined our protection PKPD outcome as viral coverage fraction such that we can determine what % of all viruses have protection above a certain threshold value *X*:

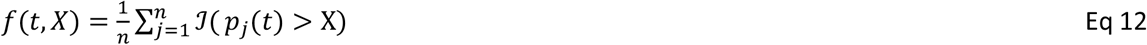

where 𝒥 is the indicator function equal to 1 if the inequality holds and 0 otherwise.

We assessed PKPD at the trough time (12-weeks, T) and as an average over the administration period (AUC/T over time through T).

## Data Availability

All simulation code is available from GitHub (https://github.com/bryanmayer) and our tool https://bnabpkpd.fredhutch.org is freely available for use.

