## Supplementary Information for "Optimizing clinical dosing of combination broadly neutralizing antibodies for HIV prevention"

### Supplementary Methods

#### Generalized titer derivation for single antibody/virus combinations

In sera, neutralization is measured using an assay typically estimating the ID50 neutralization titer: the dilution by which the sera neutralizes the *in vitro* by virus by 50% compared to a control. We define the general neutralization titer, $\text{ID}_T$, representing the dilution factor applied to sera to achieve $T$ neutralization. That is, the concentration that would have to be multipled by a factor of $\delta_T$, where

$$\delta_T = 1/ID_T \qquad \text{Eq S1}$$

to achieve $T$ neutralization. Using the Hill equation for neutralization $\nu_{ij}(t)$, see **Eq 2** in the main text, we define $V_T$ as the neutralization probability for a given target titer. For example, for the ID50 titer, we are targeting $V_{50}$ = 0.5. From there, we have the following equation:

$$V_T = \frac{1}{1+\left(\frac{IC_{50}}{\delta_T C}\right)^h} = \frac{1}{1+\left(ID_T\frac{IC_{50}}{C}\right)^h} \qquad \text{Eq S2}$$

This is 'undiluted' neutralization. We explicitly illustrate $\delta_T$ to highlight that we are estimating a scaling factor affecting the concentration, while $ID_T$ (i.e., the reciprocal) is often reported. The remaining calculations will be performed relative to $ID_T$. For a single virus, bnab combination, the $ID_T$ can be solved as a function of $V_T$ as

$$ID_T = \frac{C}{IC_{50}}\left(\frac{V_T}{1-V_T}\right)^{-1/h} = \tau\left(\frac{V_T}{1-V_T}\right)^{-1/h} \qquad \text{Eq S3}$$

Of note, any titer can be calculated relative to the ID50 ($\tau$, see **Eq 1** in the main text) given a Hill slope; and the calculation of the ID50 not depend on the Hill slope in this formulation. For example the $\text{ID}_{80}$, another common endpoint, can be calculated as follows

$$ID_{80} = ID_{50}4^{-1/h} \qquad \text{Eq S4}$$

If the Hill-slope is 1, as suggested by **Supplementary Figure 1**, then the ID50 and ID80 are related by a constant factor of 4.

#### Estimation of Hill slope using IC50 and IC80

Whereas the generalized titer (**Eq S3**) represents a ratio of a fixed concentration to a target IC value, a generalized IC value is a concentration that achieves a target neutralization level. Denoting a target inhibitory potential as P, the solution for a general IC is derived from the Hill function (**Eq. 2** in the main text) as

$$IC_P = IC_{50}\frac{P}{1-P}^{1/h} \qquad \text{Eq S5}$$

For the IC80, we then have

$$IC_{80} = IC_{50} * 4^{1/h}, \qquad \text{Eq S6}$$

and the Hill slope can be subsequently calculated,

$$h = \frac{\log 4}{\log\left(^{IC_{80}}/_{IC_{50}}\right)}. \qquad \text{Eq S7}$$

**Derivation of theoretical combination titers**

We defined a combination titer (ID$_T$) as a dilution factor applied to equivalently to all antibodies that reduces neutralization to a target level. That is, the combination titer derived here is the predicted experimental titer applied to sera containing a mixture of combined antibody concentrations.

*Additivity titer derivation*

In this derivation, we consider a combination titer across multiple antibodies, denoted *i*, against a single virus. The additivity interaction for neutralization is defined as follows:

$$v^{add} = 1 - \left(1 + \sum_i \frac{c_i}{IC50_i}\right)^{-1}. \qquad \text{Eq S8}$$

Similar to Eq S2, we incorporate ID$_T$ as follows,

$$V_T = 1 - (1 + \sum_i \frac{c_i}{ID_T IC50_i})^{-1}. \qquad \text{Eq S9}$$

We can separate out the individual bNAb titers ($\tau_i$, **Eq 1** in main text) and re-arrange such that

$$\frac{V_T}{1-V_T} ID_T = \sum_i \tau_i, \qquad \text{Eq S10}$$

and the solution follows:

$$ID_T = (\frac{V_T}{1-V_T})^{-1} \sum_i \tau_i. \qquad \text{Eq S11}$$

The additivity ID50 titer follows,

$$ID_{50}^{add} = \sum_i \tau_i. \qquad \text{Eq S12}$$

*Bliss-Hill independence titer derivation*

The Bliss-Hill independence interaction against a single virus is formulated as following for Hill functions, $v$,

$$v^{BH} = 1 - \prod_i (1 - v_i) \qquad \text{Eq S13}$$

Again, similar to Eq S2, we incorporate ID$_T$ as follows,

$$V_T^{BH} = 1 - \prod_i \frac{1}{1+\frac{c_i}{ID_T IC50_i}} \qquad \text{Eq S14}$$

For simplicity, we will explicitly solve the Bliss-Hill titer for two products against a single virus (not indexed) with a Hill slope = 1 as there is a closed formulation. We start by re-writing out the BH formulation for the generalized ($\mathrm{ID}_T$) in the underlying Hill functions. That can be rearranged as the following equation

$$\frac{V_T}{1-V_T} = \left(\frac{1}{ID_T}\right)^2 \tau_1\tau_2 + \frac{1}{ID_T}(\tau_1 + \tau_2). \qquad \text{Eq S15}$$

expressed relative to the reciprocal of $ID_T$ identifying a quadratic relationship. The solution follows:

$$ID_T = \frac{2\tau_1\tau_2}{-(\tau_1+\tau_2)\pm D} \qquad \text{Eq S16}$$

where D is the discriminant:

$$D = \sqrt{(\tau_1 + \tau_2)^2 + 4\tau_1\tau_2 \frac{V_T}{1-V_T}}. \qquad \text{Eq S17}$$

Note that while there are theoretically two solutions (as $D \geq 0$ when either $\tau$ is positive), the solution using the negative of the discriminant is outside of biological observation and we consider only the positive solution.

For an $\mathrm{ID}_{50}$ titer, we find:

$$ID_{50}^{BH} = \frac{2\tau_1\tau_2}{-(\tau_1+\tau_2)+\sqrt{(\tau_1+\tau_2)^2+4\tau_1\tau_2}}. \qquad \text{Eq S18}$$

Solution for combinations of greater than 2 antibodies can be similarly derived into polynomial equations of reciprocal $ID_T$, but may not have closed-form solutions. For example, for 3 antibodies, a cubic equation emerges

$$\frac{V_T}{1-V_T} = \left(\frac{1}{ID_T}\right)^3 \tau_1\tau_2\tau_3 + \left(\frac{1}{ID_T}\right)^2 (\tau_1\tau_2 + \tau_1\tau_3 + \tau_2\tau_3) + \frac{1}{ID_T}(\tau_1 + \tau_2 + \tau_3). \qquad \text{Eq S19}$$

but a numerical solution is required.

**General IIP with Hill slopes not equal to 1**

The full IIP calculation depends on the Hill slope as follows:

$$\mathrm{IIP}_{ij}(t) = -\log_{10}\left[1 - \nu_{ij}\left(t,\ h_{ij}\right)\right] = \log_{10}\left[1 + \tau_{ij}(t)^{h_{ij}}\right]. \qquad \text{Eq S20}$$

Assuming the Hill slope is 1, the critical inputs are time (concentration by-proxy) and IC50. Subsequently, the ID50 titer is then sufficient to calculate IIP for a given virus using single bNAb or in combinations (with the exception if BH as discussed in the next section). To calculate IIP using other IC inputs, they can be converted to IC50 using **Eq S5**.  For example, incorporating **Eq. S6**, IIP can be calculated using input concentrations relative to IC80 as follows

$$\mathrm{IIP}_{ij}(t) = \log_{10}\left[1 + 4\left(\frac{C_i(t)}{\mathrm{IC80}_{ij}}\right)^{h_{ij}}\right] = \log_{10}\left[1 + 4 * \mathrm{ID80}_{ij}(t)^{h_{ij}}\right]. \qquad \text{Eq S21}$$

As discussion in the next section, this formula for IIP applies to single bNAb calculations or calculations under the minimum, maximum, and additive interactions. IIP can also be calculated using only IC50 and IC80 by using **Eq S7** to substitute out the Hill slope, but that formulation is not simply expressed.

**Bliss-Hill independence combination titer and combination IIP**

*Derivation: BH pooled titer does not uniquely predict neutralization-derived IIP*
In this section, we will ignore time and consider the IIP definition for an individual antibody, *i*, neutralizing a single virus (omitting the virus index):

$$\mathrm{IIP}_i = -\log_{10}[1-\nu_i] = \log_{10}[1+\tau_i]. \qquad \text{Eq S22}$$

For combination titers or neutralization (**Table 1**), the following relationship holds for three of the interactions, $k \in$ [add, min, max]:

$$IIP^{(k)} = -\log_{10}\left[1-\nu^{(k)}\right] = \log_{10}\left[1+\tau^{(k)}\right]. \qquad \text{Eq S23}$$

where (k) denotes a combination measure. Briefly, we will prove this equivalence for the additivity interaction. The additivity titer formula is derived above:

$$\tau^{(add)} = \sum_i \tau_i, \qquad \text{Eq S24}$$

with

$$\nu^{(add)} = 1-[1+\sum_i \tau_i]^{-1} = 1-\left[1+\tau^{(add)}\right]^{-1}. \qquad \text{Eq S25}$$

From here, we can show that calculating IIP through combination neutralization or with combination titer is equivalent via **Eq S23**:

$$IIP^{(add)} = -\log_{10}\left[1-\nu^{(add)}\right] = -\log_{10}\left[1-(1-\left[1+\tau^{(add)}\right]^{-1})\right] = \log_{10}\left[1+\tau^{(add)}\right] = IIP^{(add)} \qquad \text{Eq S26}$$

Subsequently, an additive combination titer has a bijective relationship with IIP: a combination additive titer uniquely maps to an IIP—even when calculated using a neutralization definition—and vice versa.

The combined IIP and combination titer bijective relationship does not hold for the Bliss-Hill interaction if IIP is calculated from the neutralization definition. Starting with the combination neutralization definition, denoted by superscript (BH, *v*), we find:

$$IIP^{(BH,\nu)} - \log_{10}\left[1-\nu^{(BH)}\right] = -\log_{10}[1-(1-\prod_i (1-\nu_i))] = -\sum_i \log_{10}[1-\nu_i], \qquad \text{Eq S27}$$

effectively the sum independent IIPs via **Eq S22**. However, if consider the combination titer derived version of IIP, denoted (B$H,\tau$), we fin

$$IIP^{(BH,\tau)} = \log_{10}\left[1+\tau^{(BH)}\right] \qquad \text{Eq S28}$$

and the relationship suggested via **Eq S23** fails to hold

$$-\sum_i \log_{10}[1-\nu_i] = \sum_i \log_{10}[1+\tau_i] \neq \log_{10}\left[1+\tau^{(BH)}\right]. \qquad \text{Eq S29}$$

Forgoing formal proof, we consider a simple example where $\tau_1 = \tau_2 = 9$. Calculating BH IIP via neutralization results (**Eq S27**) in an IIP = 2 whereas calculating BH IIP via combination titer (**Eq S28** via **Eq S18**) results in an IIP = 1.36. In fact, there are infinite positive combinations $\{\tau_1 = x, \tau_2 = \frac{10^2}{x+1} - 1\}$ that generate an $IIP^{(BH,\nu)} = 2$ and maps to different $IIP^{(BH,\tau)}$.

There is a mechanistic interpretation when calculating IIP using the combination neutralization derivation (**Eq S27**): the Bliss-Hill interaction is applied at the event-level first where a single combined neutralization estimate is generated and then translated through the IIP definition. The interpretation using the combination titer is different and effectively a distance metric: the combination titer is a factor applied to all concentrations that adjusts the current combined neutralization to 50%. The interpretation of this metric on the IIP-scale (**Eq S28**) is not immediately clear, especially as it is not mathematically equivalent to the neutralization definition assuming using Bliss-Hill independence (**Eq S29)**. Generally, an IIP calculated from combined BH titer appears to be lower than when calculated from combined neutralization (See next section). Interestingly, the combined IIP can be equivalently calculated using either combination neutralization or the combination titer for the additivity, maximum, and minimum interactions; so an IIP can be interpreted both ways under those assumptions.

*Implications: BH pooled titer does not uniquely predict neutralization-derived IIP*

Here, we illustrate a simple, practical example, assessing combined neutralization of two bNAbs at varying fixed total concentrations, as in a dosing context. We compared performance of two potential concentration ratios (1:1 or 1:10) against a single virus (IC50 of 1 and 0.1 concentration units, respectively). First, we calculated the predicted experimental combination titer (**Table 1**) under additivity and Bliss-Hill independence (**Supplementary Figure 3A**). The predicted titers are relatively similar under each interaction model, both predicting higher titers from the 1:10 ratio across the range of concentrations. Next, we calculated the predicted neutralization IIP (**Eqs S25 and 27**) for each interaction model across total concentrations (**Supplementary Figure 3B)**.  The conclusions for additivity remain the same as with titer; however, under Bliss-Hill, the conclusions changes: at higher concentration the Bliss-Hill model predicts higher neutralization using the 1:1 ratio. This is consistent with the mechanism of the Bliss-Hill interaction model, specifically rewarding more balanced ratios as the virus must escape all products independently.

Lastly, we mapped the combination titer to neutralization IIP in this example and compared to the expected relationship from single virus/antibody combinations depicted in **Eq 3** (**Supplementary Figure 3C**). For additivity, all lines overlap indicating a consistent, coherent relationship between titer and neutralization as proven above in **Eq S26**. This was not the case for Bliss-Hill, where deviation in the lines indicate that the combination titer does not uniquely predict neutralization. For example, at a Bliss-Hill combination titer of 100, the 1:1 ratio elicits the highest neutralization, and both ratios elicit higher neutralization than predicted by using titer in **Eq 3** (black line). That is, under Bliss-Hill interaction, an experimental titer measured in sera with combined bNAbs does not predict neutralization. To predict BH neutralization, the individual bNAb concentrations and viral IC50s are required.

This result has additional implications when considering correlates and protection. If a single Ab trial suggests a titer correlate, that must be translated carefully into the Bliss-Hill framework. Specifically, as done in the empirical case study in the main manuscript, we derive independent protection estimates for each bNAbs first then apply the Bliss-Hill independence model to get a combined protection estimate. This is in contrast to an alternative strategy of calculating a combination titer first then calculating protection from the dose-response relationship depicted in **Supplementary Figure 4C**. As suggested in **Supplementary Figure 3**, this approach could underestimate true protection. Similar issues may arise from applying “potency reduction factors” at the combination titer-level rather than at each the individual bNAb-level first.

**Supplementary Tables and Figures**

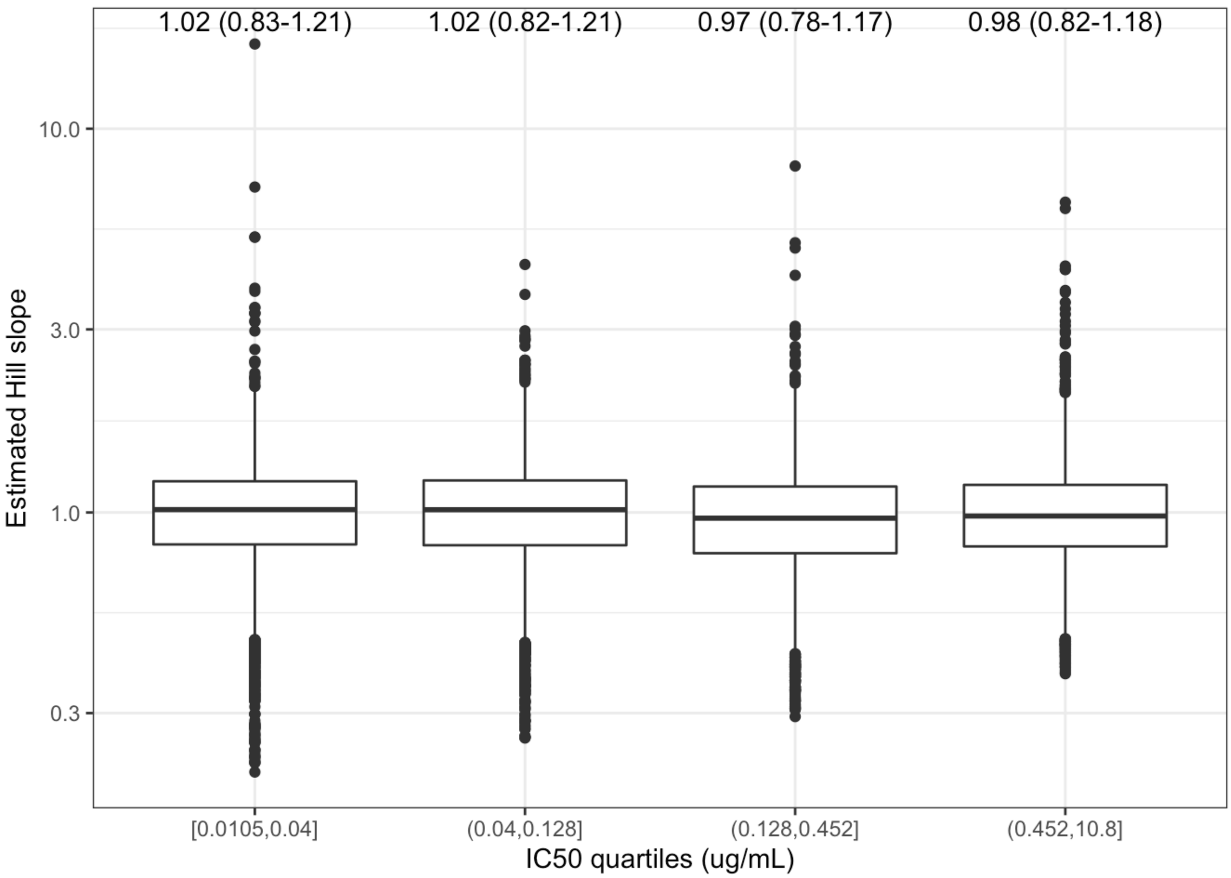


**Supplementary Figure 1. Estimated neutralization Hill slope using CATNAP data.** Estimated Hill slopes for different ranges of IC50 measurements (split into quartiles). Each point represents a calculation for an antibody/virus combination in the database where IC50 and IC80 measurements were both within 0.01-20 ug/mL range. Median Hill slope estimates (IQR) displayed above each box plot. There were approximately 5000 measurements per quartile.

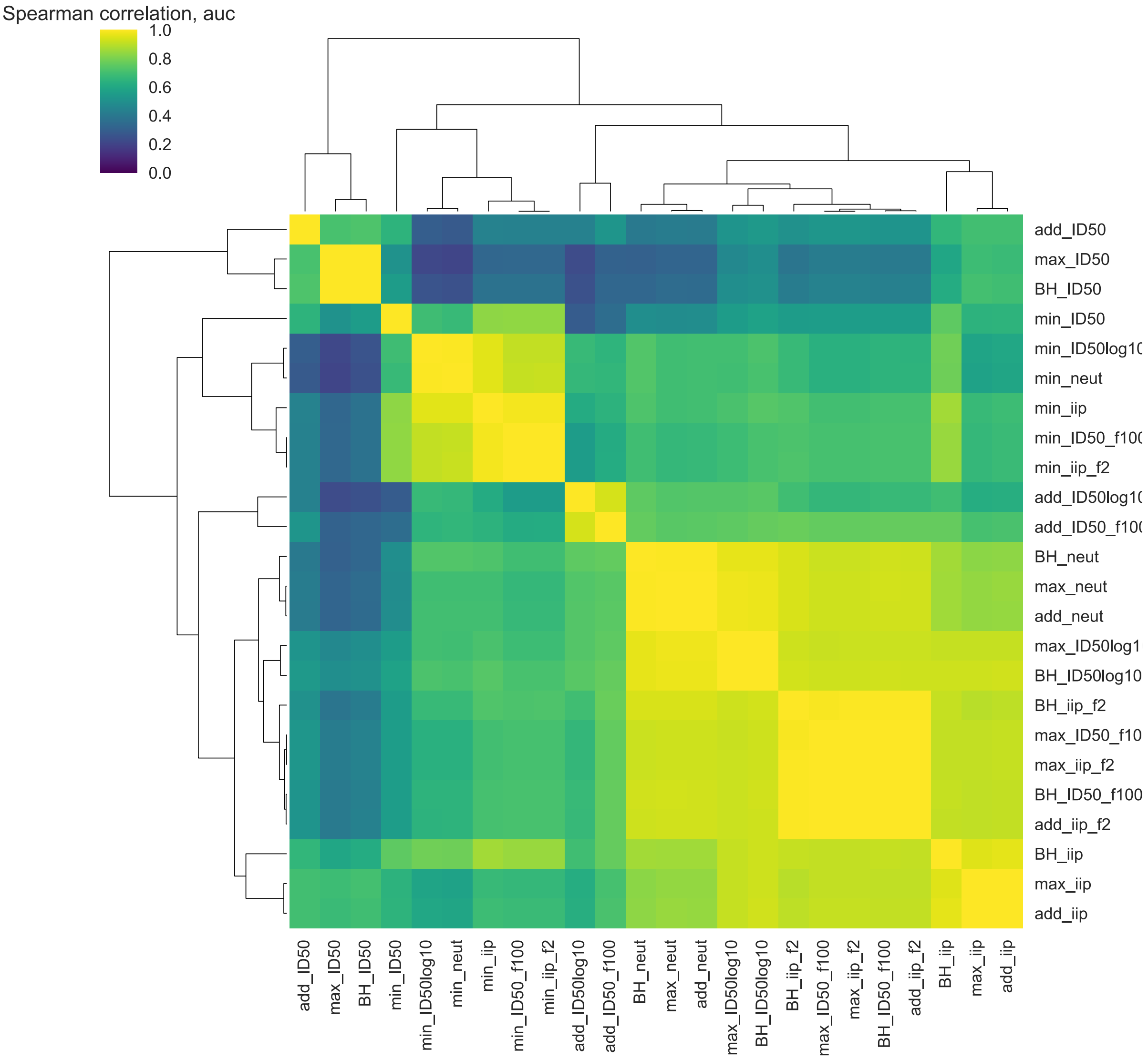


**Supplementary Figure 2. Additional clustering results for AUC.** As for trough in **Fig 2A**, endpoints cluster by Spearman correlation into similar 5 main categories, from top to bottom: titer, minimum, additive titer, neutralization/coverage, and IIP.

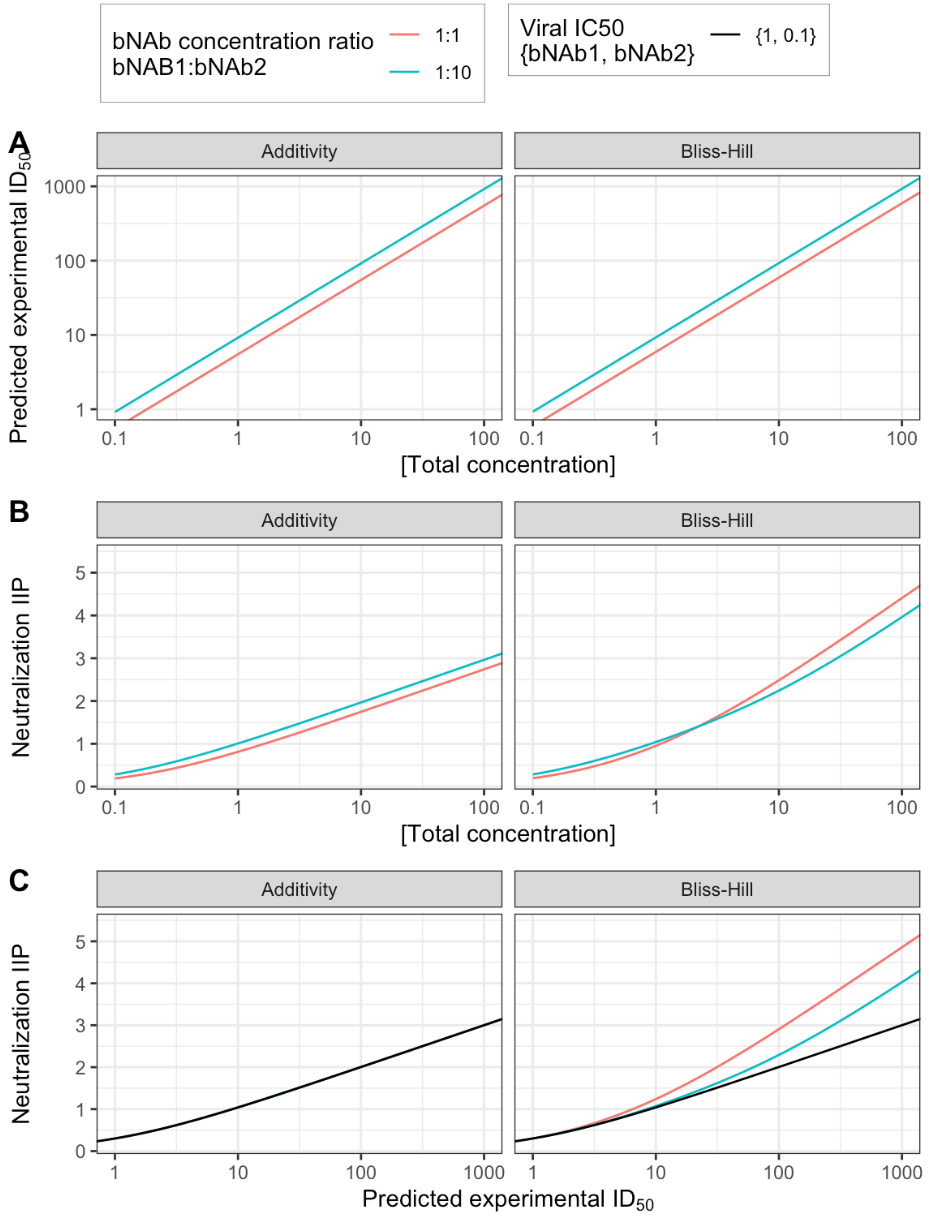


**Supplementary Figure 3. IIP and ID50 relationship by interaction model against a single virus at varying concentration and bNAb ratios. A)** The predicted combination titer by interaction model (see **Table 1** for $ID_{50}$ titer formulas) across total bNAb concentrations at two ratios of the individual bNAb (1:1 and 1:10 denoted by colors). **B)** The predicted neutralization IIP by interaction model (see **Table 1** and for neutralization formulas) across total bNAb concentrations and ratios. IIP was calculated as the $log_{10}$-transformation of one minus the combined neutralization (**Eq 3**). For Bliss-Hill, the lines cross at increasing concentration indicating the 1:1 ratio performs better, a qualitatively different conclusion than **A**. **C)** Relationship between predicted combination ID50 titer and neutralization across total bNAb concentrations and ratios. The black line indicates the predicted relationship between IIP and titer for single antibody/virus combinations (**Eq 3**). For additivity, all lines overlap indicating the relationship holds. For Bliss-Hill, separate lines indicate that the combination titer does not correspond to a unique neutralization. For example, at a BH combination titer of 100, the 1:1 ratio elicits the highest neutralization, and both ratios elicit higher neutralization than predicted by using titer in **Eq 3**.

**Table S1:** Population PK input parameters for the empirical case study optimization. The -T variants derived to extend half-life based on PK analysis of parental variants.

| parameter | 10-1074-T | 3BNC117-T | VRC07-523LS |
|---|---|---|---|
| Vc | 4.50 | 5.02 | 1.89 |
| Cl[a] | 0.09 | 0.22 | 0.09 |
| Q | 0.67 | 2.19 | 0.41 |
| Vp | 3.29 | 8.65 | 2.34 |
| ka[b] | 0.39 | 0.39 | 0.39 |
| bioavailability[b] | 0.46 | 0.46 | 0.46 |
| HL (days) | 64.57 | 45.62 | 35.76 |

[a]Clearance parameter derived for -T variants by dividing study-estimated clearance by three-fold to extend the half-life.
[b]Parameter data only available from [PMID: 31473167].

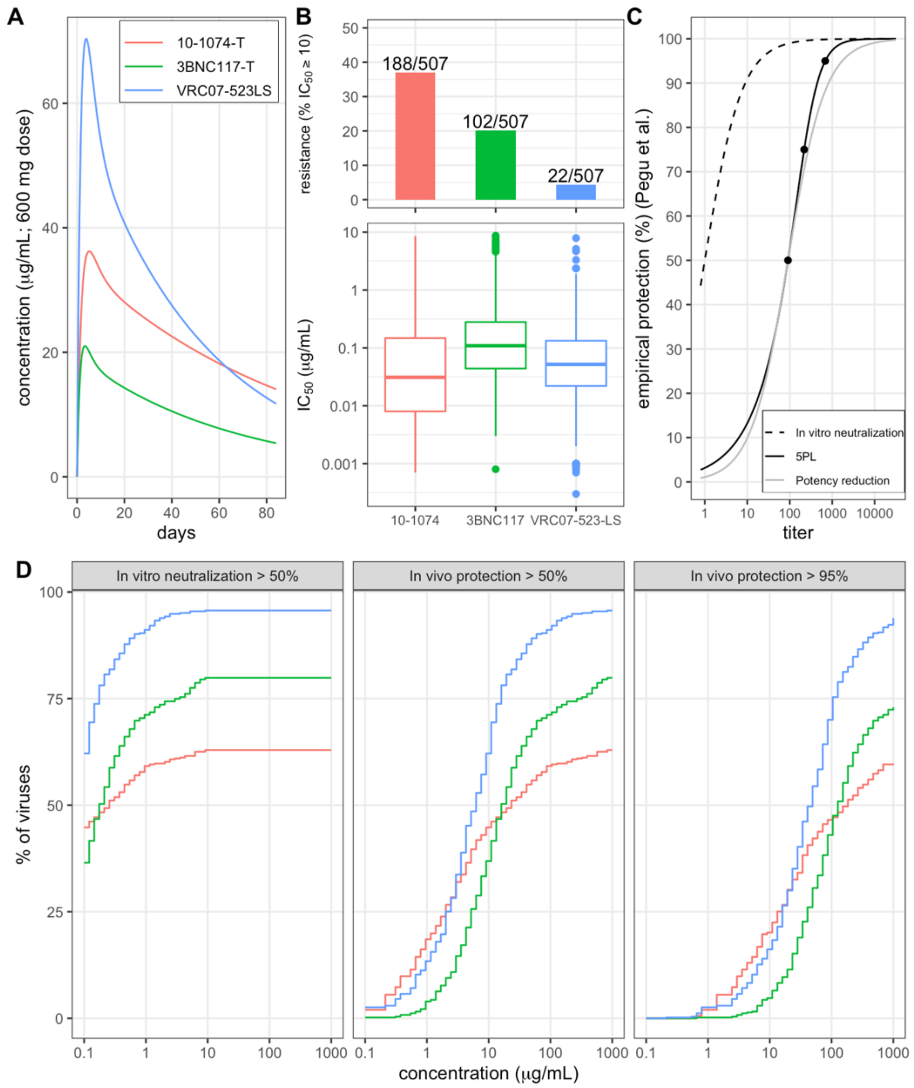


**Supplementary Figure 4. Input data for 3-bnab combination optimization.** A) PK over 12 weeks for each of the products given at a 600mg dose using subcutaneous route. B) Neutralization data for each of the products from 507 viruses in CATNAP database. Distribution of IC50s among sensitive viruses (IC50 < 10) are shown as box plots in the bottom plot and barplots depict the total resistant viruses (IC50 > 10) in the top plot. C) Relationship between titer (concentration divided by IC50) and *in vivo* protection using the Pegu et al. NHP challenge meta-analysis for single bNAb administration. Expected *in vitro* neutralization at given titer shown as dashed line, potency reduction depicted curve shift achieving 50% protection in Pegu *et al.* shown as gray line, and 5PL model curve fitted over three Pegu *et al.* protection estimates (points at 50%, 75%, and 95% protection) depicted via black line. D) For increasing concentrations for the individual products, the percent of viruses sensitive at given thresholds (facets) and concentrations (x-axis). The first facet depicts neutralization coverage when concentration exceeds the IC50 values depicted in **B**. The second two facets depict protection coverage comparing titer (given concentration over IC50) to 50% (second panel) and 95% (third panel) 5PL protection thresholds in **C**.

**Table S2: Ratio optimization results from 3-bNAb optimization for 3BNC117-T, 10-1074-T, and VRC07-523-LS. See Methods for details.**

| | | | Optimal dosing proportion | | |
|---|---|---|---|---|---|
| **% protection target** | **Time endpoint** | **% viral coverage** | **3BNC117-T** | **10-1074-T** | **VRC07-523-LS** |
| 50% | AUC | 88 | 0.04 | 0.19 | 0.77 |
| | Trough | 78 | 0.07 | 0.20 | 0.72 |
| 95% | AUC | 49 | 0.01 | 0.27 | 0.72 |
| | Trough | 31 | 0.08 | 0.33 | 0.59 |
